# The Canadian Collaborative Project on Genetic Susceptibility to Multiple Sclerosis cohort population structure and disease etiology

**DOI:** 10.1101/2024.04.18.24305992

**Authors:** Alison M. Pagalilauan, Elif Everest, Suzanna Rachimi, Daniel Reich, Alex D. Waldman, A. Dessa Sadovnick, Carles Vilariño-Guell, Michael J. Lenardo

**Author notes:** Corresponding author (MJL).

## Abstract

**Background:** Previous genetic and epidemiological studies have examined subpopulations from the Canadian Collaborative Project on Genetic Susceptibility to Multiple Sclerosis (CCPGSMS) patient cohort, but an encompassing analysis of the study population has not yet been carried out.

**Objective:** This study examines patterns of multiple sclerosis (MS) prevalence in 13,663 cohort members, including 4,821 patients with MS or suspected MS and 8,842 family members.

**Methods:** We grouped participants into epidemiologic subgroups based on age of MS onset, clinical stage at diagnosis, symptom type at disease onset, sex, proband status, disability as measured by the EDSS, and ancestry based on reported ethnicity.

**Results:** We observed a 2.7:1 MS prevalence ratio of women to men, though disease severity was greater for male patients. Variation in the age of disease onset between patients was only slightly associated with sex and strongly associated with disease type. Specific types of clinical symptoms at disease onset were associated with the prognosis. Regional residence did not correlate with disease onset, type, or severity.

**Conclusion:** Population trends, as presented here, are not explained by environmental factors alone, highlighting the need for a comprehensive genetic analysis to understand disease variance across families.

## Introduction

Multiple sclerosis (MS) is the leading cause of non-traumatic brain injury in young adults and is characterized by immune-mediated damage to the myelin nerve sheath and central nervous system (CNS) degeneration [1]. MS has varying forms and stages of the disease, including relapsing-remitting MS (RRMS, ∼85% of cases), secondary-progressive MS (SPMS, ∼60–70% of RRMS cases develop into this form), and primary-progressive MS (PPMS, ∼10–15% of cases) [2,3]. Many patients first present with an acute “clinically isolated syndrome” (CIS). If the patient’s initial magnetic resonance imaging (MRI) demonstrates CNS lesions, CIS will likely progress to relapsing disease [4]. MS pathogenesis is believed to involve environmental and genetic factors [5,6]. While the increasing prevalence of MS may be due to changes in diagnostic criteria and improvements in disease awareness in health systems around the world [7], alterations in environmental factors may also play a role. Distance from the equator, vitamin D deficiency, and infection with Epstein-Barr virus (EBV) are significant MS susceptibility factors [8,9]. In the United States, the highest MS prevalence is in northeastern and midwestern states, generally at higher latitudes [2]. Prevalence in Manitoba, Canada, is among the highest worldwide [10,11]. The correlation between higher latitudes and increased risk of MS may involve decreased exposure to solar ultraviolet (UV) radiation [12]. Reduced exposure to UV radiation was first proposed as an environmental susceptibility factor for MS to explain how vitamin D protects from MS [12]. Vitamin D controls the promoter region of the human leukocyte antigen (HLA) allele *HLA-DRB1*, the main susceptibility locus for MS [13]. The specific haplotype is *HLA-DRB1*1501*, dominant in Northern European populations [13]. Like MS, EBV prevalence is also increased at higher latitudes and a risk factor for MS [14,15]. However, the specific pathogenic mechanisms of environmental factors are unknown, leading to interest in defining genetic factors.

The most strongly associated genetic risk factors identified with MS are certain HLA II genes, such as DQA1, DQB1, and DRB1 [16,17]. Genome-wide association studies (GWASs) have identified a host of genes, mainly immunological, that may contribute to MS [18]. How these genes contribute to MS pathogenesis remains unknown. Given the persisting questions regarding MS etiology and pathogenesis, it is valuable to examine historical cohorts using modern epidemiological techniques and genomic analyses. One classic study of MS is the Canadian Collaborative Project on Genetic Susceptibility to Multiple Sclerosis (CCPGSMS), for which information is still publicly available [5,21]. A total of 13,663 patients and family members were included before enrollment ended. Exome sequencing analyses have been performed for a subset of CCPGSMS families, revealing candidate risk variants associated with MS [19,20]. Here, we provide information on population structure and disease etiology of the complete CCPGSMS cohort.

## Methods

This study is a retrospective analysis of data assembled by the Canadian Collaborative Study Group. The investigation uncovered MS patients with pediatric onset of disease, families with multiplex MS disease, and MS co-occurring with other autoimmune diseases. Participants provided written informed consent under an IRB protocol approved by the Research Ethics Review Board (REB # H08-01669) at the University of British Columbia. Data were collected over 18 years, providing a large patient and family member cohort with thorough background information [21]. Enrollment ended approximately 10 years ago. Genetic screens, geography questionnaires, and co-morbidity questionnaires were administered to all patients.

When the study first began in 1993, patients were diagnosed using the Poser criteria. These now obsolete criteria required evidence of two or more relapses lasting over 24 hours, occurring a minimum of one month apart, in addition to evidence of CNS lesions [22]. Where possible, patients and family members were later re-evaluated using the McDonald criteria, which requires evidence of CNS damage over time [23,24]. Patients and their families were categorized as “affected,” “unaffected, blood-related,” and “unaffected, married-in” based on disease status and relationship to the proband.

The investigators of the present study did not collect the original specimens or clinical information, which were de-identified for use in the present study. NIH reviewed the study and determined an IRB-approved protocol was not necessary because it did not meet the basic requirements to be considered human subject research. Data collected during the original CCPGSMS study were further divided by age of MS onset (i.e., age at date of diagnosis for clinical symptoms), clinical stage at diagnosis, symptom type at disease onset, sex, proband status, disability as measured by the EDSS, and ancestry based on reported ethnicity.

The mean ages of onset in different groups of patients were compared using the Kruskal-Wallis test. The differences in the types of symptoms at disease onset in different patient groups that were defined based on disability scores and clinical stages were tested using the chi-squared test for trend. All statistical analyses were performed with R (v4.2.1) in RStudio (v2023.06.1). The package ms.sev (v1.0.4) was used to perform global age-related MS severity (ARMSS) score calculations [25]. We used ggplot2 (v3.3.6) and plotrix (v3.8.2) to generate plots and display the results of statistical tests. The distribution of age at MS onset was analyzed in 10-year increments (Table 1; Fig 1). Using cancensus [26] (v0.5.5) and tidycensus (v1.4.4), we mapped the probands across Canadian provinces. The package cancensus allowed us to access Canadian census data available via Statistics Canada (2022).

**Fig 1.**
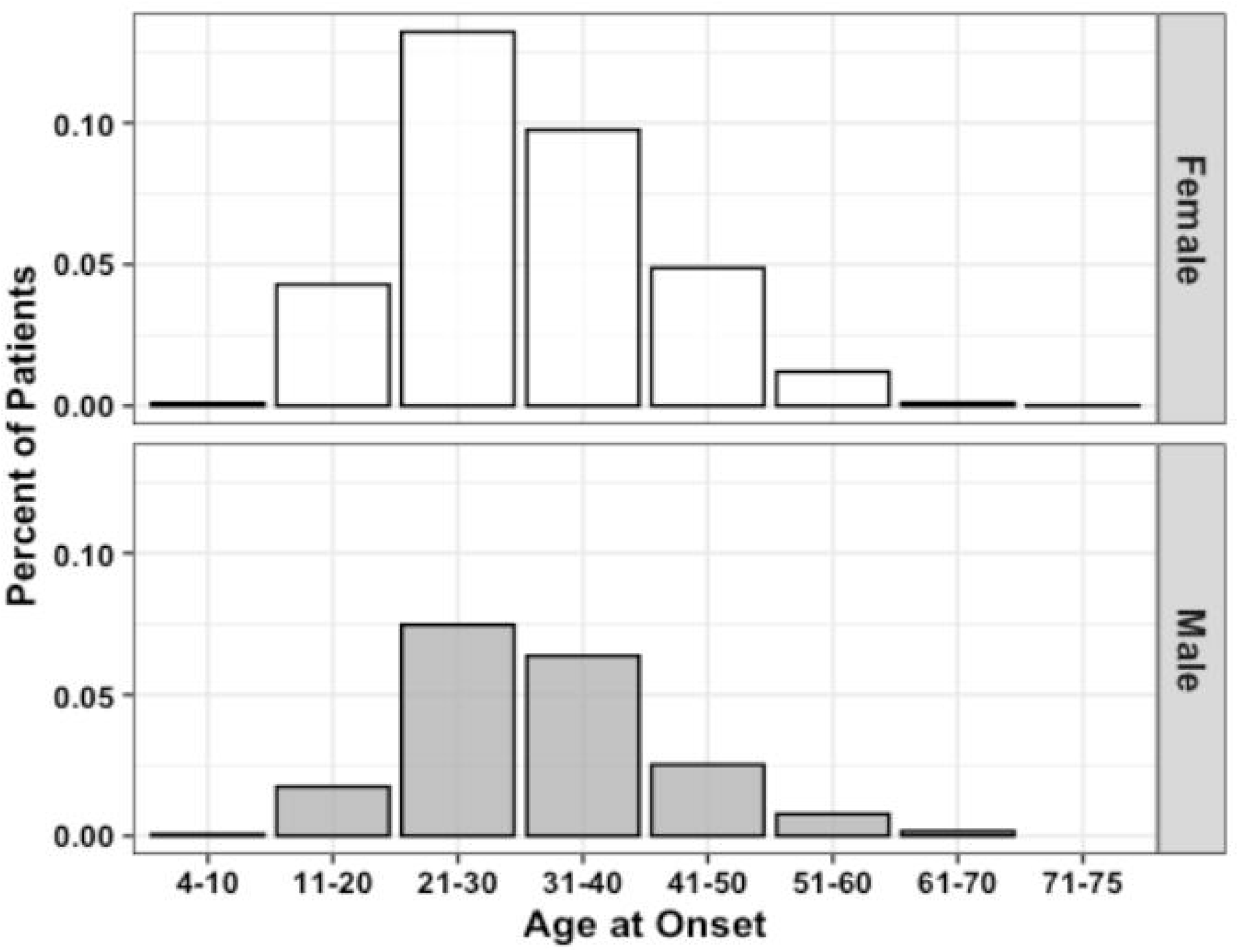
Age of MS onset distribution by sex (n=3,787). Data represent all patients for whom age at initial diagnosis was provided.

**Table 1.**
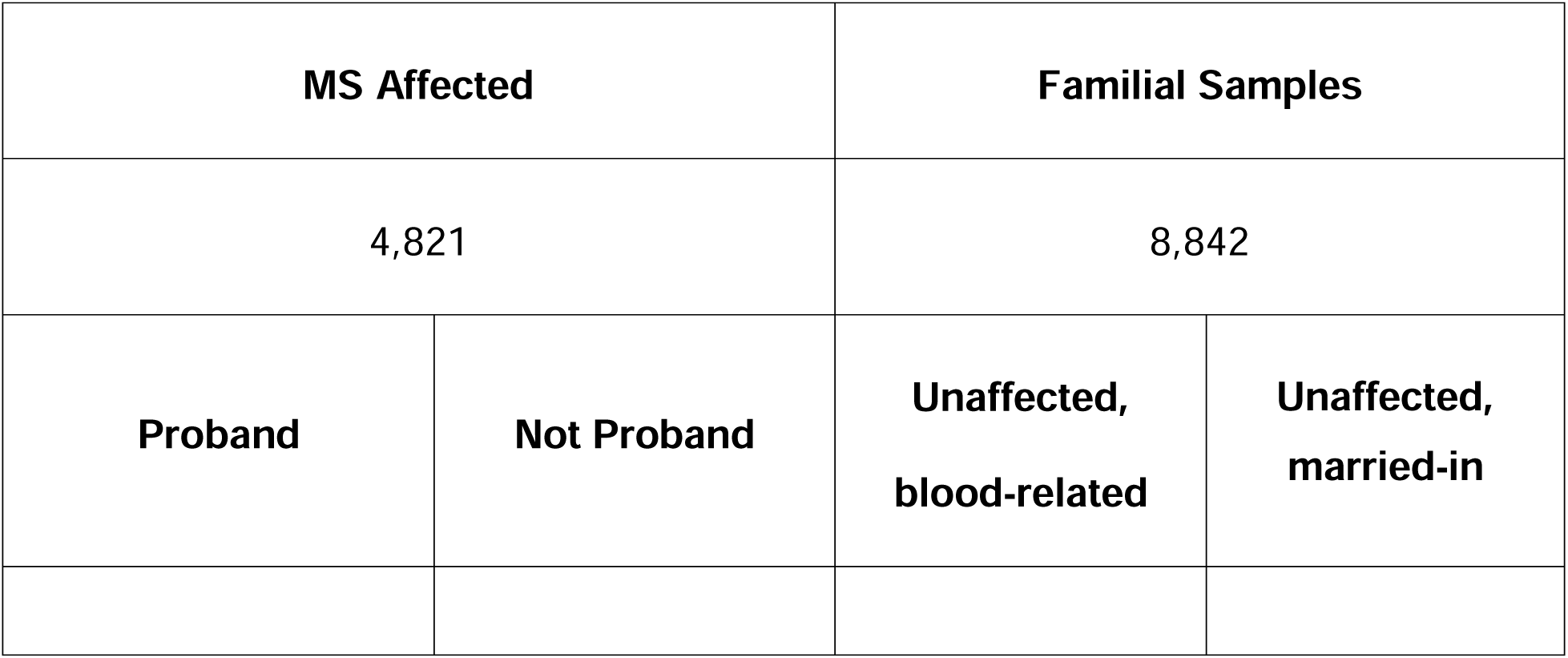
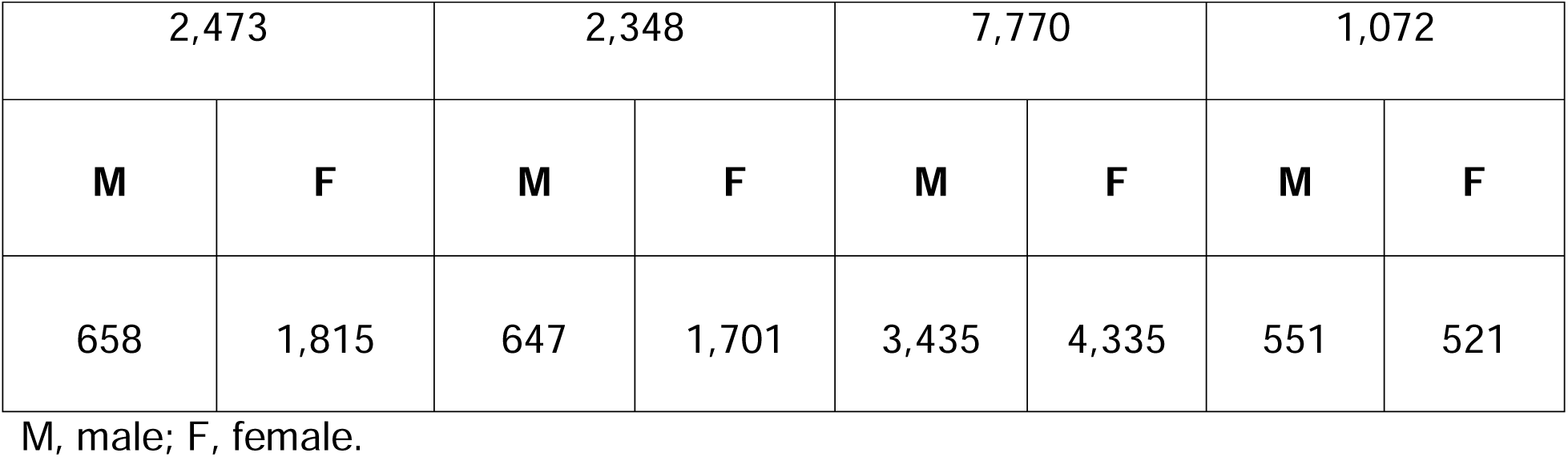
Summary of population data included in the study, subdivided based on disease status, blood-relation or marriage status, proband status, and sex (n=13,663).

| MS Affected |  | Familial Samples |  |
| --- | --- | --- | --- |
| 4,821 |  | 8,842 |  |
| Proband | Not Proband | Unaffected,<br>blood-related | Unaffected,<br>married-in |

|  |  |  |  |  |  |  |  |
| --- | --- | --- | --- | --- | --- | --- | --- |
| 2,473 |  | 2,348 |  | 7,770 |  | 1,072 |  |
| **M** | **F** | **M** | **F** | **M** | **F** | **M** | **F** |
| 658 | 1,815 | 647 | 1,701 | 3,435 | 4,335 | 551 | 521 |

M, male; F, female.

EDSS scores were converted to ARMSS scores [25]. This disability measure normalizes EDSS scores relative to disease duration to detect disability differences between different age groups. We used an ARMSS-based disability metric instead of the MS Severity Score (based on disease duration), as well as the EDSS (snapshot disability indicator), to represent disease severity status better since disease duration information can be incorrect due to the possibility of subclinical MS stages with varying periods. ARMSS scores were only calculated for patients with known age at EDSS collection available (n=1,746). ARMSS scores were used to generate global ARMSS (gARMSS) scores for each patient using the age of data collection and EDSS data. Because sample sizes were not equal between the male and female proband groups, a Monte Carlo simulation was used to randomly sample 475 patients from the female proband group over a series of 10,000 iterations with the male population and calculate an average gARMSS within a sample size equal to that of the male proband group (n=475).

## Results

The population data gathered by the CCPGSMS represented a total of 13,663 individuals: 4,821 patients with MS (approximately 35%), subdivided into probands (51%) and affected relatives (49%), and 8,842 unaffected family members (Table 1). Patients included male=27% and female=73%. Unaffected family members were grouped by blood relation (88%) or marital status (12%) and again divided by sex (male=45%, female=55%) (Table 1). We found that although the sex ratio of unaffected individuals was comparable (females/males=1.2), the affected individuals were skewed towards women (females/males=2.7). We next examined patterns of MS incidence (Table 2; Fig 1), which was highest among individuals 21 to 30 years old, independent of sex (Fig 1), followed by those aged 31 to 40 years. MS was extremely rare under the age of 10 (n=7) and over 60 (n=19), probably due to lower awareness of early- and late-onset MS at the time of the sample collection period.

**Table 2.** Proportion of the affected population grouped by age at the time of MS.

| **Age Stratum** | ***n*** | **Proportion (%)** | **Cumulative Proportion (%)** |
| --- | --- | --- | --- |
| **4–10** | 11 | 0.29 | 0.29 |
| **11–20** | 449 | 11.86 | 12.15 |
| **21–30** | 1490 | 39.35 | 51.50 |
| **31–40** | 1147 | 30.29 | 81.78 |
| **41–50** | 532 | 14.05 | 95.83 |
| **51–60** | 139 | 3.67 | 99.50 |
| **61–70** | 18 | 0.50 | 99.80 |
| **71–75** | 1 | 0.03 | 100 |

RRMS was the most common clinical form (34.8%), followed by chronic-progressive MS (CPMS) (10.4%) and relapsing-progressive MS (RPMS) (7.5%). Benign diagnoses describing a less severe version of RRMS with few mild relapses were grouped with the larger RRMS group. Though “chronic progressive” is no longer commonly used, we believe it is important to reflect the cohort demographics of the original study. We also did not have enough clinical information to associate CPMS data with more specific categories such as SPMS or PPMS. Patients with benign phenotypes were defined as having an EDSS score equal to or lower than 3.0 after 15 or more years from disease onset and having never received MS therapies [19,27]. SPMS (4.1%) and PPMS (1.7%) were the least frequently reported clinical forms. Affected patient diagnoses that were unreported at the time of data collection were considered “unknown” (41.5%) (Fig 2A). Disease type contributed strongly to the variation in age of onset between patients. Mean age of onset of patients with a progressive disease was significantly higher than that of patients with benign and relapsing forms of MS (both p<0.0001).

**Fig 2.**
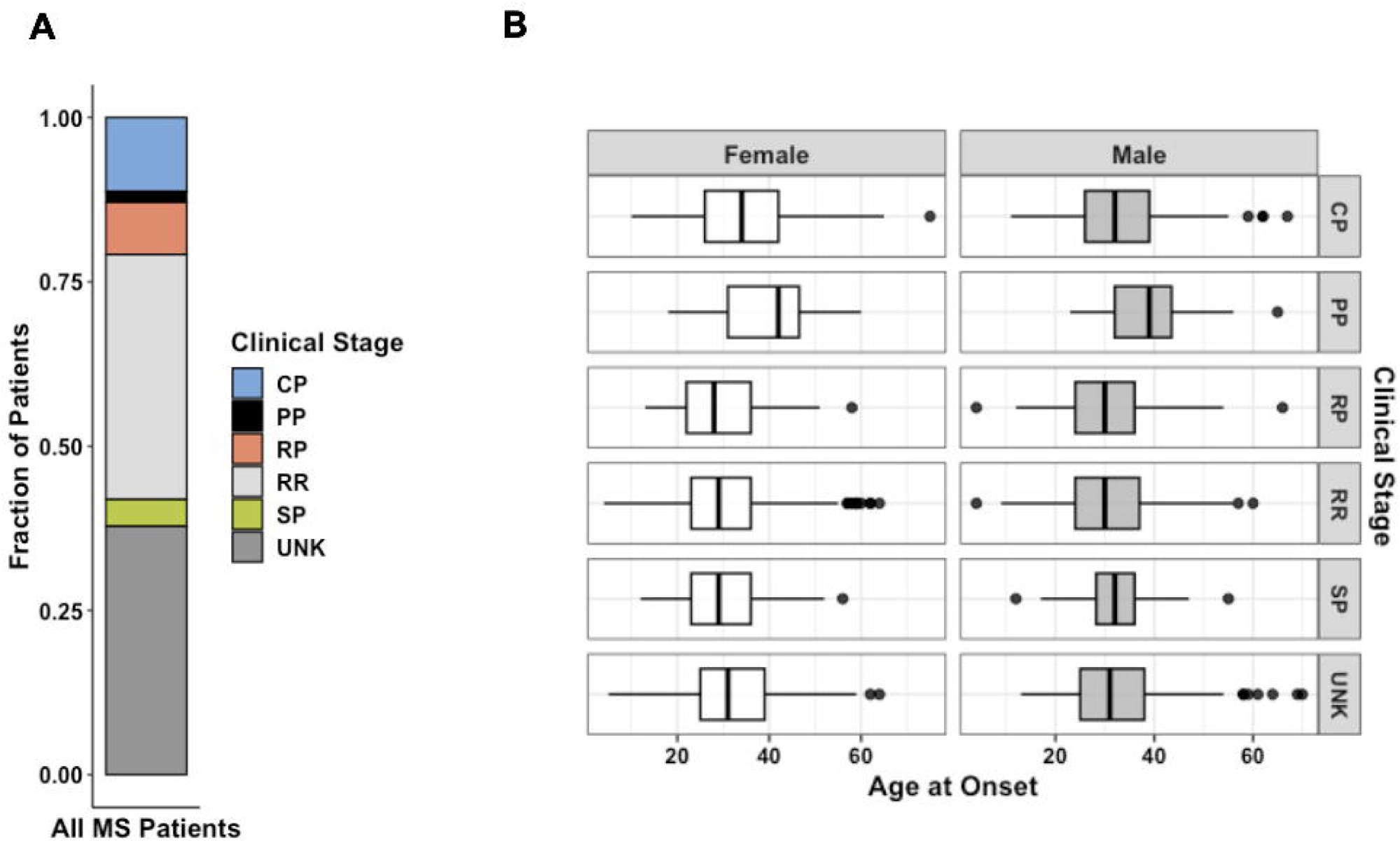
(A) Distribution of MS forms for all patients diagnosed as affected from the patient cohort (n=4,821). (B) Faceted boxplots characterizing age of onset by sex and MS clinical stage at the time of data collection (n=3,787). Data represent all patients for whom age at initial diagnosis was provided. CP, chronic-progressive; PP, primary-progressive; RP, relapsing-progressive; RR, relapsing-remitting; SP, secondary-progressive; UNK, Unknown.

The ancestry breakdown was examined for proband patients for whom data were available (n=905). Most participants reported ancestry from Europe (92.5%), far exceeding those reporting ancestry from Asia (5.7%), Africa (1%), and Indigenous North America (0.05%) (Fig 3). Reported ancestry did not significantly contributed to variation in age of onset (*p*=0.06).

**Fig 3.**
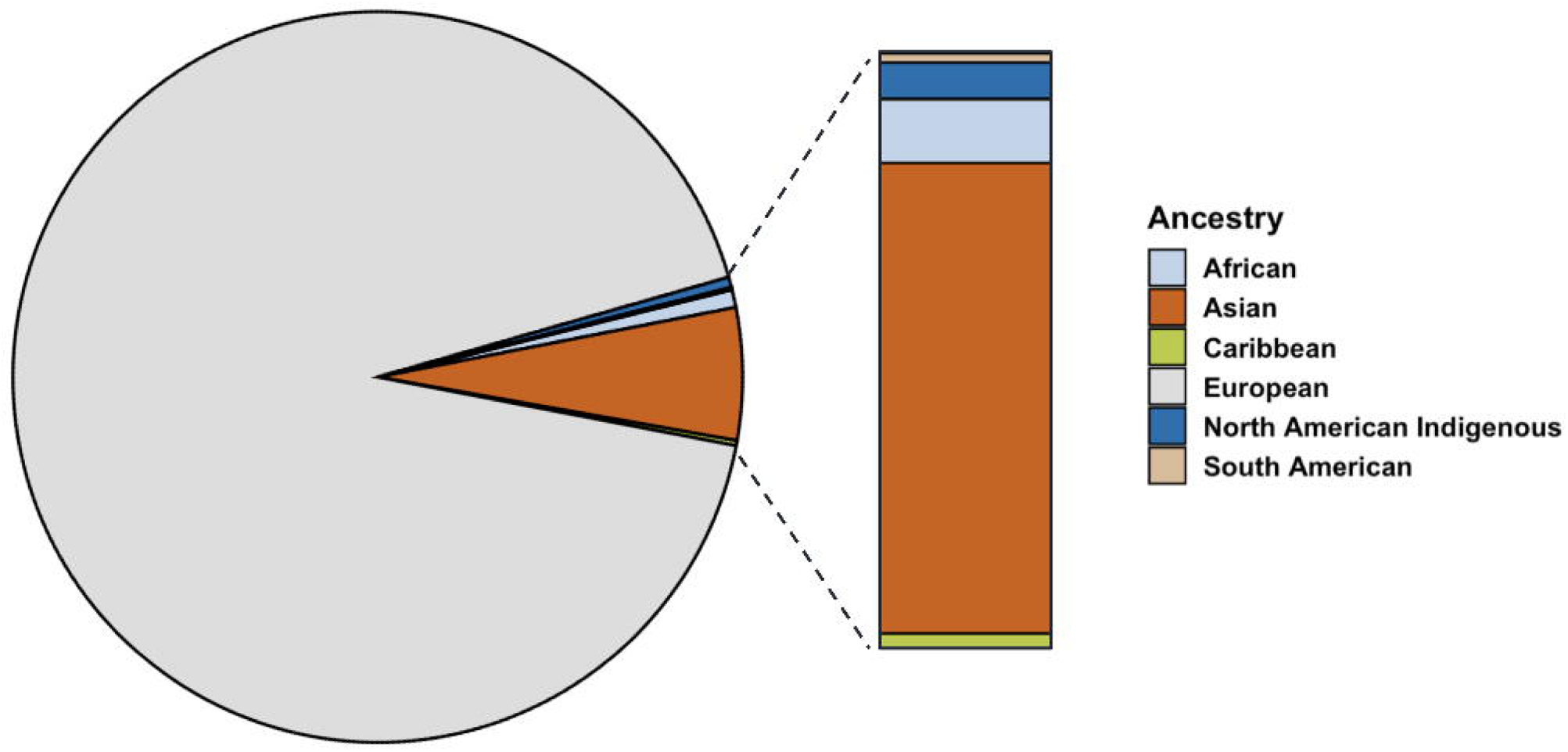
Distribution of proband patients grouped by reported ancestry (n=905).

Residency at the time of data collection was also examined and plotted by province to show the distribution of proband patients across Canada as a dot density map (Fig 4A). The total population per province, as estimated by the 1996 Census, was indicated on the same dot density map to highlight the influence of geographic distribution on overall cohort demographics. Patients were mainly from Ontario (33.2%) and Quebec (17.7%), the most populous provinces, followed by British Columbia (15.2%), Nova Scotia (10.8%), and Saskatchewan (10.6%) (Fig 4A). There appeared to be an ascertainment bias towards patients recruited in British Columbia and Ontario, as the study was spearheaded in parallel by MS clinics in Vancouver and London, though we observed enrichments in probands (individuals with familial MS) in Nova Scotia and Saskatchewan (Fig 4B). Provincial residency did not appear to contribute significantly to variation in onset age.

**Fig 4.**
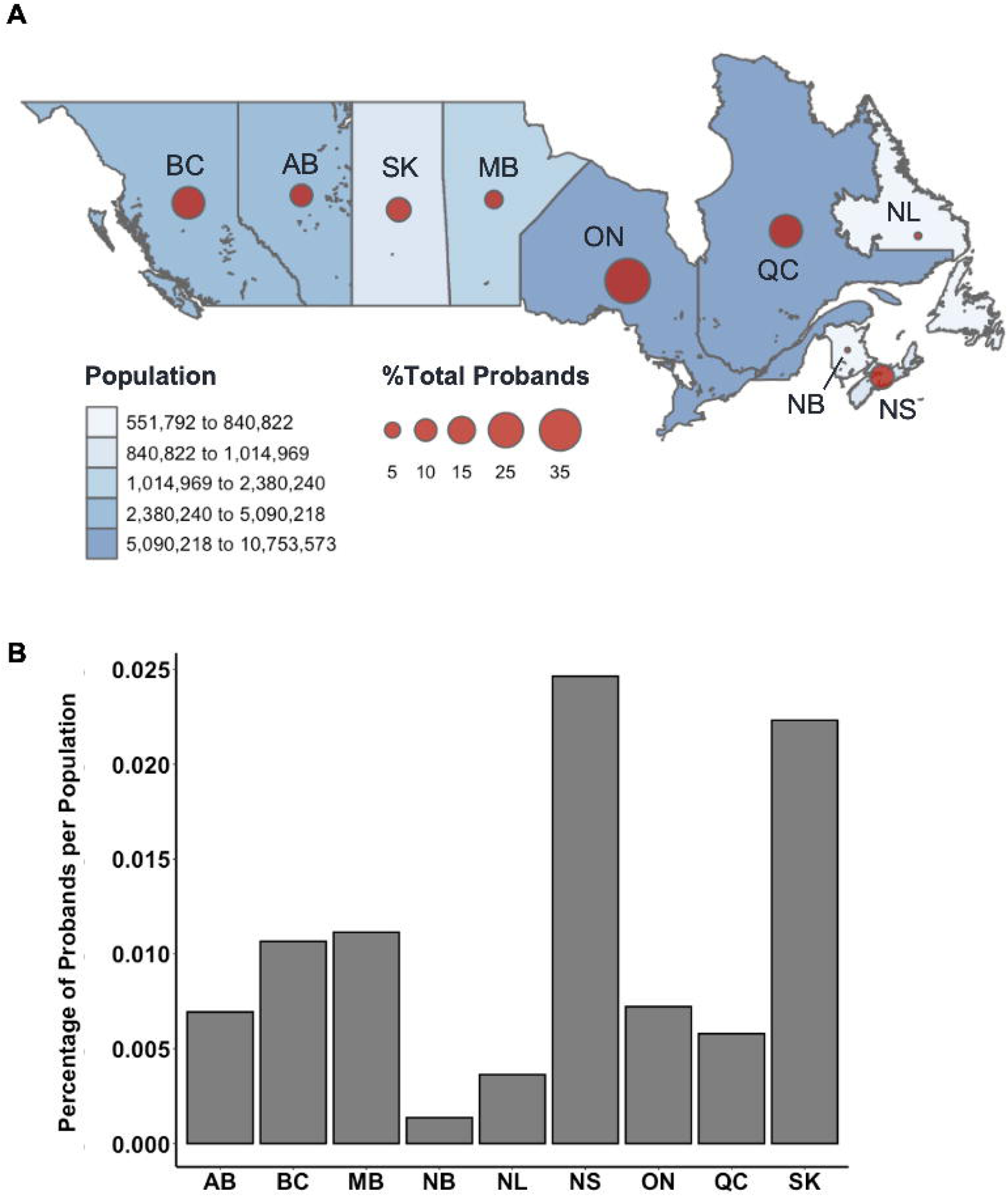
(A) Relative percentage distribution of proband patients by Canadian province (n=2,372). Datapoints are proportionally sized to the relative percentage of probands and overlayed with total population per province as estimated by the 1996 Census (accessed via Statistics Canada). Probands from unknown provinces (n=101) were excluded from this distribution. (B) Relative percentage of MS proband patients per province compared to the relative percentage of the total Canadian population per province. BC, British Columbia; AB, Alberta; SK, Saskatchewan; MB, Manitoba; ON, Ontario; QC, Quebec; NL, Newfoundland; NB, New Brunswick; NS, Nova Scotia.

To interpret differences between subsets of patients with MS, disability status was examined using ARMSS scores. Affected males had significantly higher gARMSS scores compared to affected females (*p*=0.00095) (Fig 5A). To account for the discrepancy in sample size between male and female proband subsets, this sex-biased difference in disease severity was further corroborated by a probabilistic model with a Monte Carlo simulation generating 10,000 simulated data sets built from female proband EDSS data equaling the sample size of the male proband EDSS data (n=475). The mean ARMSS of each simulated data set was compared to the mean gARMSS score of the male proband group (*p*<2.2×10^-16^). Across all iterations of the simulation, the mean difference in gARMSS scores between groups remained small but constant, with male proband scores maintaining an average 0.53 increase over female proband scores (Fig 5B).

**Fig 5.**
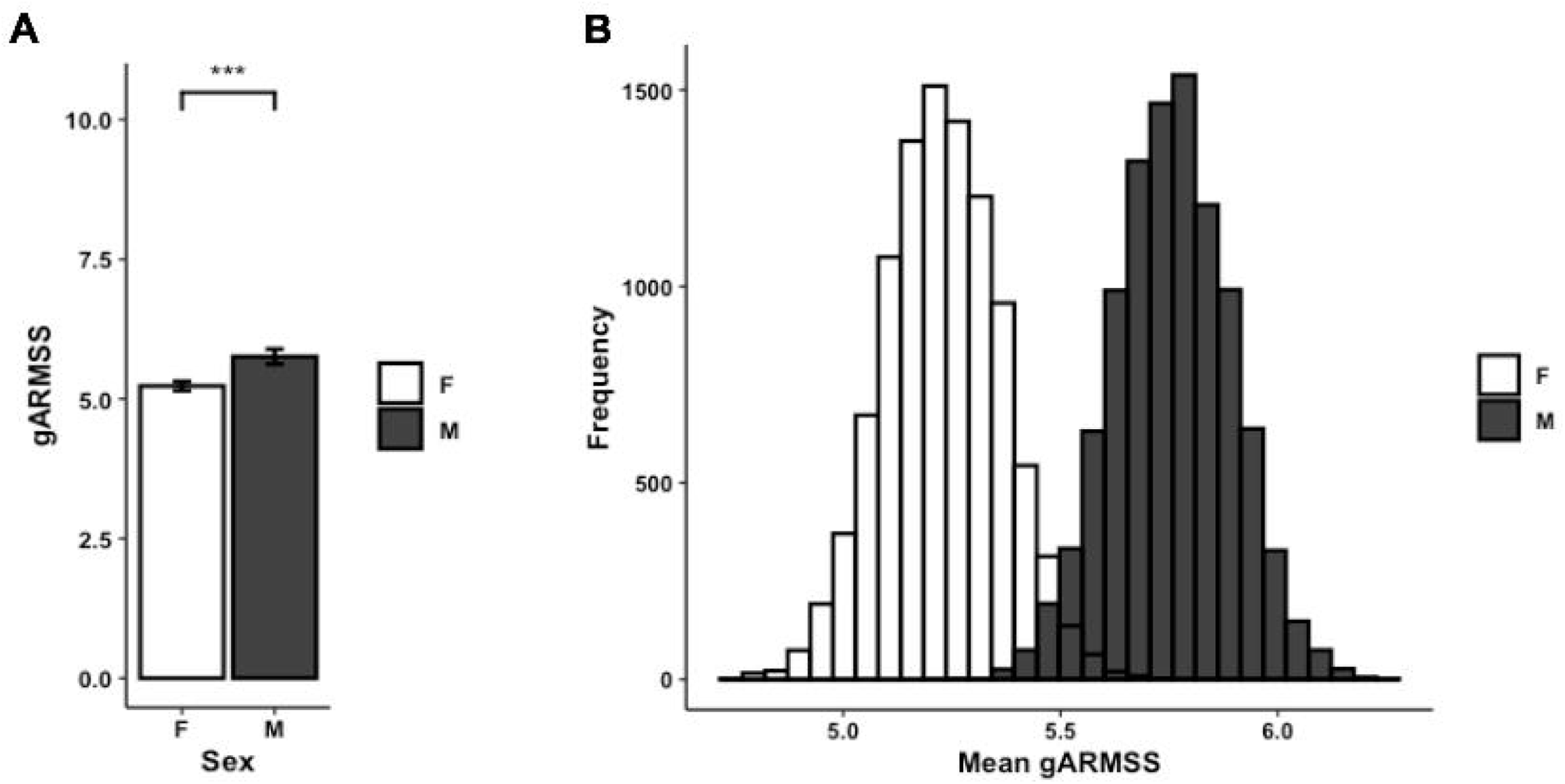
(A) Average gARMSS of proband patients by sex with error bars indicating standard of the mean (n=1,746; p=0.00095). Data represent all proband patients for whom EDSS at initial diagnosis was provided. Two-tailed paired Student t-test p-value indicate statistical significance (***, p<0.001). (B) Histogram of gARMSS means of proband patients by sex, randomly sampled by Monte Carlo simulation with 10,000 iterations to equalize sample size. EDSS, Expanded Disability Status Scale; gARMSS, global age-related MS severity.

To assess associations between symptoms at disease onset and prognosis, patients with available gARMSS scores were stratified into two groups: those with a gARMSS score ≥5 (indicative of an unfavorable disease course, n=1,264) and those with a gARMSS score <5 (indicative of a favorable disease course, n=495). We used this cutoff based on a previous study indicating that the cutoff well-distinguished the patients based on their disability statuses [28]. Indeed, the mean gARMSS score in the current patient cohort was significantly higher in the unfavorable disease course group (*p*<0.0001), showing that the disability scores of each group were not closely aggregated below and above the cut-off value of 5. It is important to note that the higher number of patients with a gARMSS score ≥5 than those with a gARMSS score <5 is likely due to the long follow-up period (>18 years) of the cohort, given that the development of a progressive course is likely an age-related phenomenon [29]. The symptoms at disease onset were categorized as follows: fatigue, diplopia, retrobulbar neuritis, balance problems, gait disturbances, upper limb ataxia, sensory-face, sensory-spinal cord, pain, motor-acute, motor-slow, transverse myelitis, bladder dysfunction, Lhermitte’s sign, and vertigo. Among individuals with available data on symptoms at disease onset, the correlation between the type of symptom and disease course was examined using the chi-squared test for trend, comparing the unfavorable and favorable course groups. A significantly higher proportion of patients in the unfavorable course group presented with gait problems and slow motor dysfunction at disease onset compared with those in the favorable course group (*p*=0.0027 and *p*=0.005, respectively), while sensory spinal cord symptoms were more prevalent at disease onset in the favorable course group (*p*=0.017) (Fig 6A). Consistently, within the subgroup of patients with a well-defined disease course history (benign/relapsing-remitting with complete remission vs. definitive progressive course), a greater percentage of individuals with progressive MS exhibited balance problems (*p*=0.022), gait disturbances (*p*=0.012), and slow motor dysfunction (*p*=0.026) at disease onset. Conversely, patients with a benign course more frequently manifested sensory spinal cord symptoms as their initial disease presentation (*p*=0.04) (Fig 6B). Given the higher disease severity observed in male patients, we tested whether there was a difference between prognosis-associated symptoms at disease onset in males and females. This revealed that sensory spinal cord symptoms were significantly more frequent in females at disease onset (*p*<0.0001). Other comparisons did not show a difference in severity that could shed light on MS pathogenesis. For example, disease severity in different provinces appeared to relate mainly to the population density of different provinces (S2 Fig).

**Fig 6.**
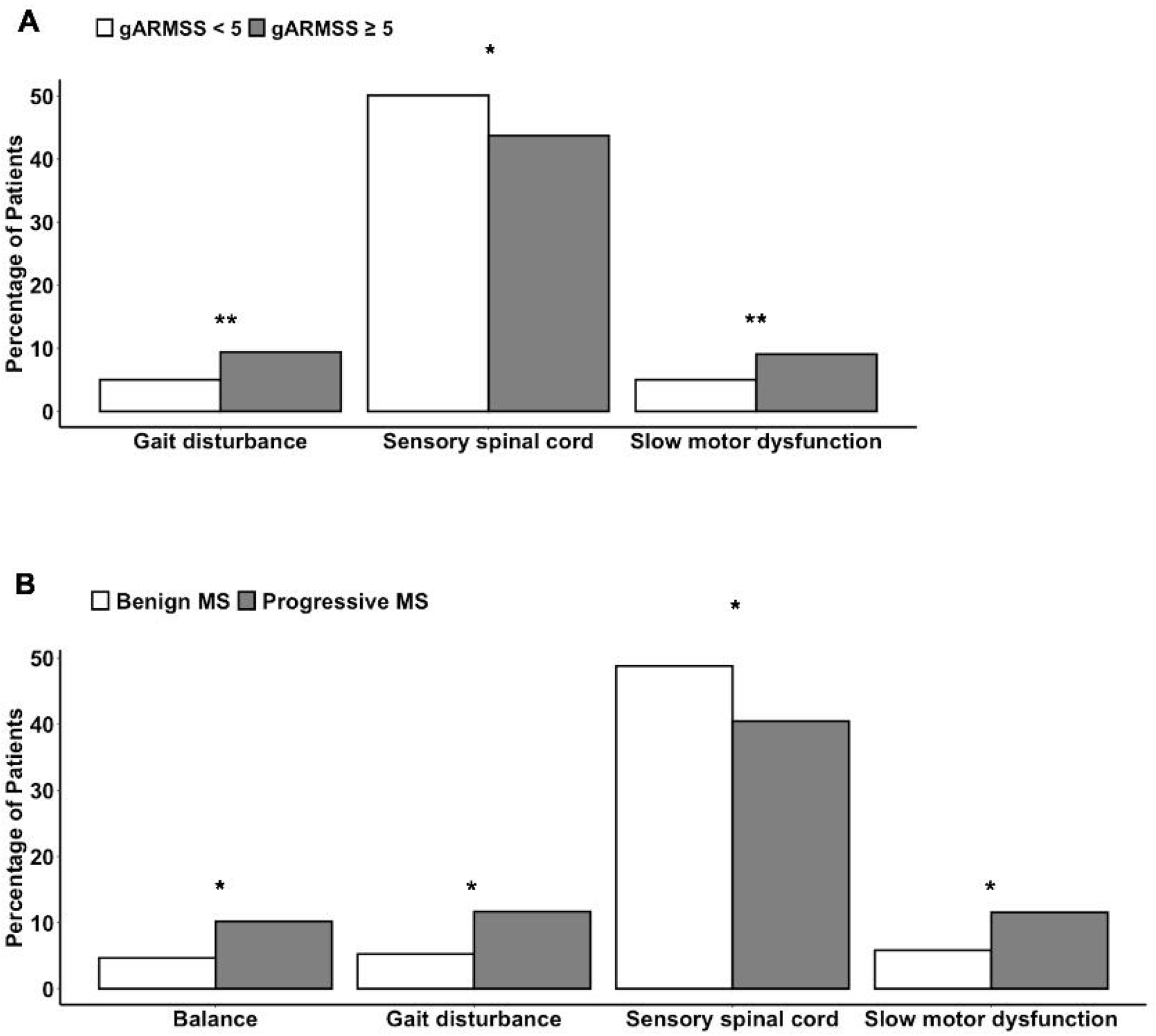
Differences in the symptoms at disease onset (A) between patients with a gARMSS score lower than 5 (favorable disease course) and those with a gARMSS score equal to higher than 5 (unfavorable disease course) and (B) between patients with a benign or progressive MS type. Chi-squared test for trend (*, p<0.05; **, p<0.01). gARMSS, global age-related MS severity.

## Discussion

Our study population of 13,663 participants comprises one of the largest familial specimen sets assembled for an autoimmune disease. Patterns of MS incidence found in this cohort corroborate those examined in previous studies. Among CCPGSMS patients, peak incidence was observed between ages 21 and 40. In both males and females, patients with CPMS and PPMS were diagnosed at slightly older ages than those with other types of MS (Fig 3B), aligning with the notion that older age predisposes patients to more progressive phenotypes or that progression reflects the natural history of the disease as patients age. We also observed that RRMS was the most common form of MS; 82% of patients in this study were younger than 41 years at the time of diagnosis, which is consistent with existing reports [15,30,31].

Over a third of proband patients reported residency in Ontario (32.7%). The remaining proband population was largely from Quebec (17.4%), British Columbia (16.7%), Nova Scotia (9.4%), and Saskatchewan (9.3%). Though the 1996 Census reported that less than 6% of the total population of Canada resided in Nova Scotia and Saskatchewan at the time, more proband patients of the CCPGSMS cohort were found to be residents of these provinces than residents of Alberta (7.9%) or Manitoba (5.2%). Studies of MS prevalence in Canada have reported the Canadian prairies (SK, AB, MB) as having the highest incidence rates of all regions [10,11]. Proband incidences in this study were proportionally comparable to the relative populations of most Canadian provinces, apart from Alberta and Nova Scotia. Notably, the relative proportion of probands in Nova Scotia was three times greater than the relative proportion of Canadians residing in the province in 1996. It is also important to note that many rural communities have reduced access to medical care, which sometimes forces affected patients to move to urban centers for better access. Conversely, some patients with increased severity may be limited in their ability to move to major centers.

Increased disease severity in males was observed in the present cohort, validating previous observations that although women with RRMS relapse more frequently than men with RRMS, the latter group exhibits more rapid disability accumulation following each relapse [32] and thus a worse overall outcome. The ratio of affected females to affected males in our study is comparable to the 3:1 ratio measured globally [33]. This demographic imbalance in sex is relatively constant across age strata in this study and has been observed in numerous other cohorts [31,32,33]. However, different hypotheses have been proposed to explain this distribution [32,34]. Regarding the differences in clinical presentation at onset, there are a limited number of studies investigating differences in attack locations between female and male patients with MS, yielding inconsistent results [35,36]. A comprehensive study involving 14,969 patients with MS examined the relationship between 49,279 phenotypically characterized MS attacks and associated demographic and clinical parameters [36], showing that sensory attacks are more commonly observed in female patients and associated with a non-progressive course. This is in line with our current observations that sensory spinal cord symptoms were more frequent in female patients and in those with a lower disability score and benign course, and male patients had higher disability scores. The reasons underlying the differences in clinical presentation and progression between sexes are currently unclear; however, cellular and animal studies have indicated possible molecular differences between sexes, involving differences in oligodendrocyte differentiation, remyelination capacity, or axonal vulnerability [37,38].

In addition to sex, ancestry is a risk factor for MS. Here, we report a skewed number of affected patients with ambiguous European ancestry, confounding risk attributable to proximity to the equator. This limitation is due to the biased geographical ascertainment in this study and the small sample size of proband patients for which complete demographic information was provided (n=905). In our analysis, the sample size was strictly limited to proband patients for which all criteria (i.e., sex, age, clinical stage, ancestry, and residence) were available. Though most enrolled participants reported European ancestry, it is unclear whether this distribution is indicative of trends unique to the etiology of MS, as ∼70% of Canadians are ethnically European [39]. This ascertainment bias is a weakness of this dataset. Regional distance from the equator was previously shown to be a risk factor in MS, but alone, it does not determine disease status. Reduced vitamin D due to more limited sunlight exposure, rather than proximity to the equator, has been discussed as the relevant correlated risk factor. In rural areas where more populations engage in outdoor work such as farming, vitamin D exposure may not always accurately correlate with latitude [40]. Confounding studies of MS prevalence in Canada have reported a higher incidence among patients from the Canadian Prairies, which are north of other Canadian regions but also experience the most average annual sunlight hours [11]. Further, many countries across the world have seen an increase in the prevalence of MS, regardless of geographical location [41]. Interestingly, more significant increases in female-to-male ratios are associated with a latitudinal gradient, with the ratio being more drastically skewed towards female patients with RRMS in northern latitudes [32,42]. Findings such as these have led many researchers to reassess the idea of a latitudinal gradient as a causative factor, further suggesting that this gradient effect has been attenuated [33].

A major limitation of the current study is that MS diagnostic criteria have changed since the data were initially collected, resulting in non-identical, multilateral patient diagnoses and severity assessments. Another limitation is that the effect of disease-modifying treatments was not accounted for in the analysis of MS severity. Lastly, there is a possible selection bias in different sub-comparisons since we could not include the whole cohort in some analyses due to missing data. The next steps for this CCPGSMS cohort will be combining the population data with genomic DNA sequencing. As multiplex MS families are common in this cohort, we plan to focus on identifying high-risk genetic variants in MS.

## Data Availability

The data that support the findings of this study are available from the corresponding author, upon reasonable request.

## Acknowledgments

We thank Justin Gabrielski for critical reading of the manuscript.

## Notes

### Competing Interest Statement

This work was supported by the Division of Intramural Research of NIAID, NIH.

### Funding Statement

This work was supported by the Division of Intramural Research of NIAID, NIH.

### Author Declarations

Research Ethics Review Board of the University of British Columbia gave ethical approval for this work (REB # H08-01669).

### Summary of Updates

Per the editor's request, we have revised Supplementary Table S1 to indicate an age range for each patient instead of the exact age.

