## Supplementary figures and images for "The Canadian Collaborative Project on Genetic Susceptibility to Multiple Sclerosis cohort population structure and disease etiology"

### Supplementary Figure 1

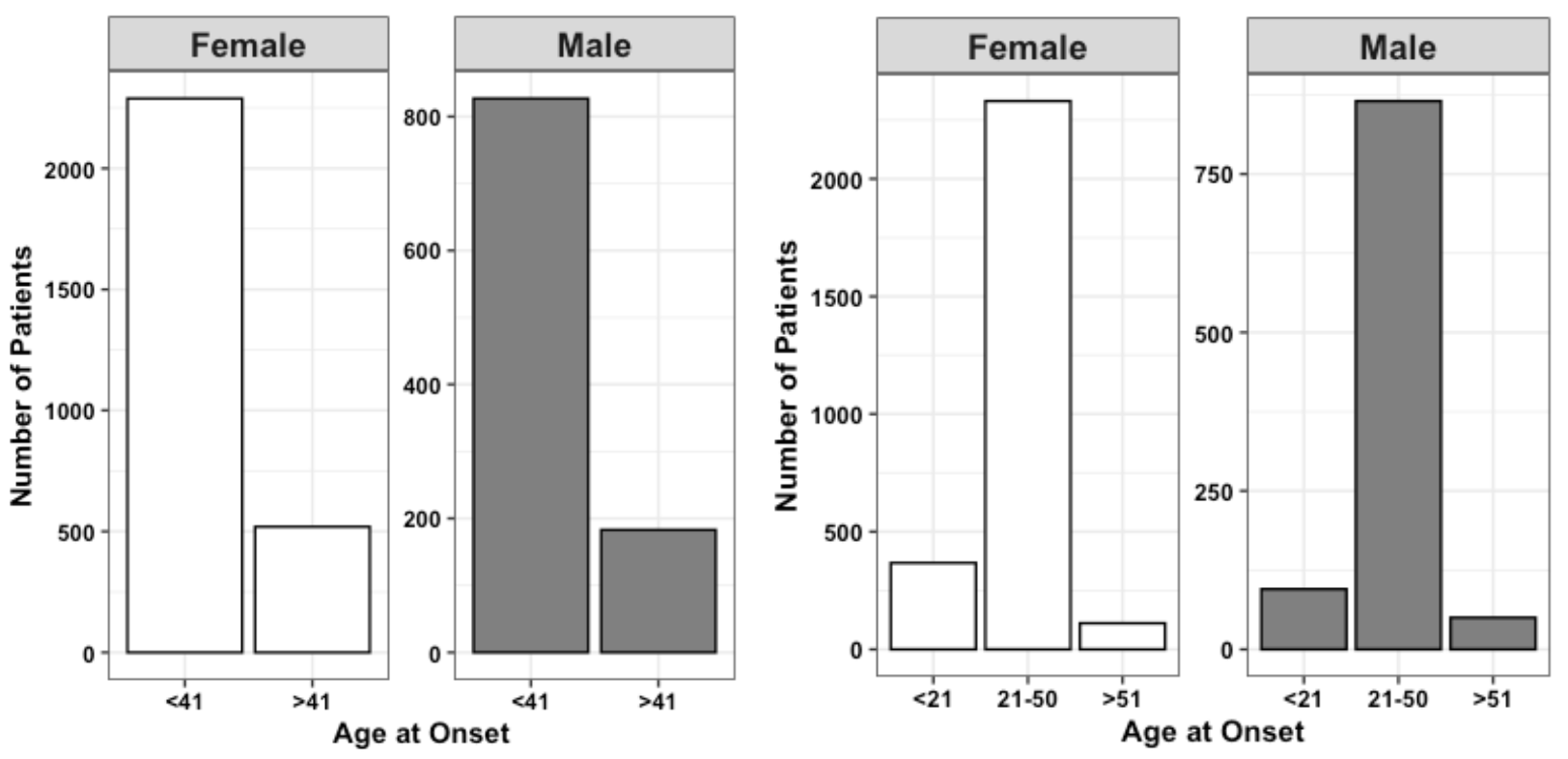

### Supplementary Figure 2

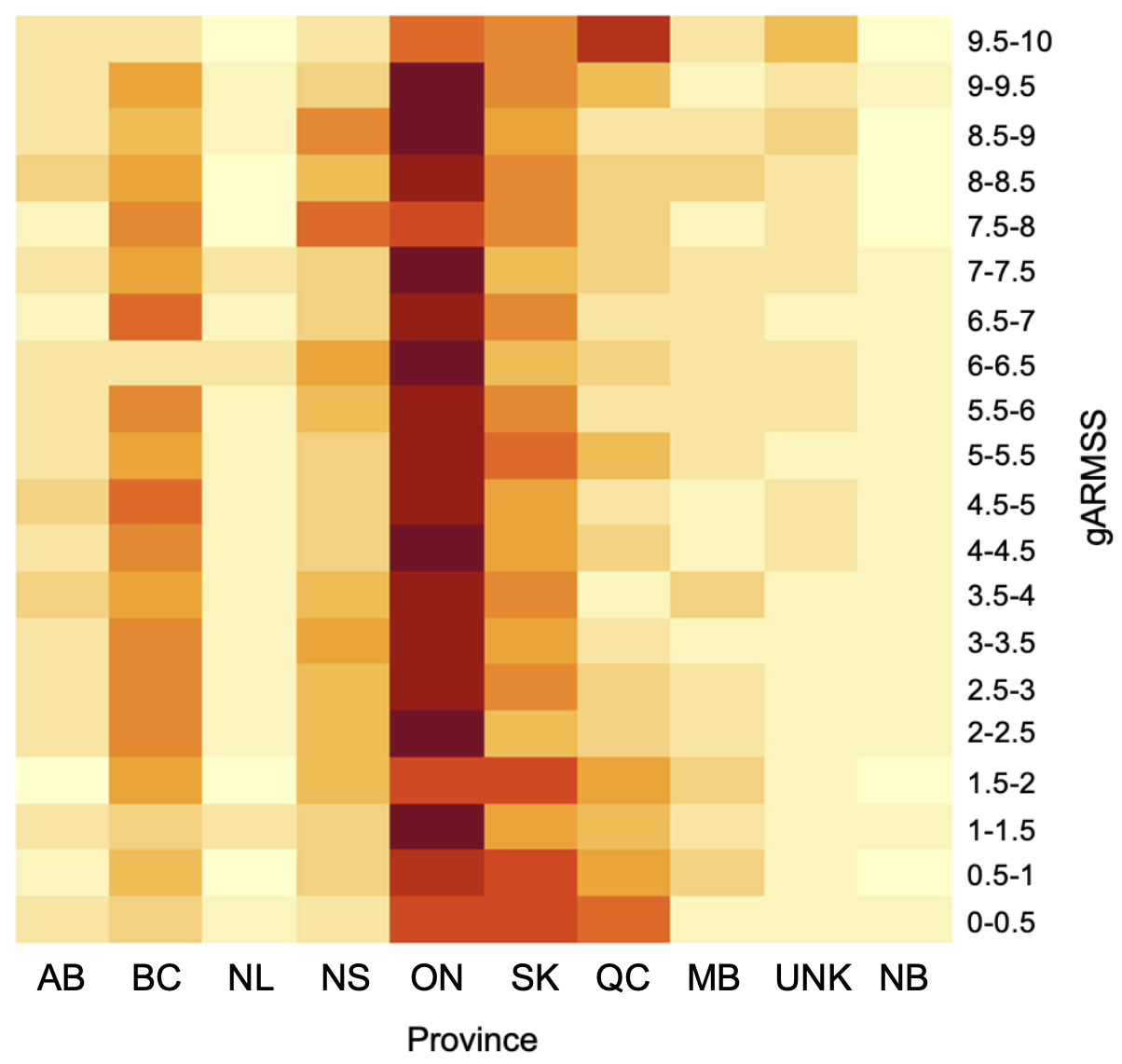
